# Characteristics and mechanism of reciprocal ST-segment depression in acute ST segment elevation myocardial infarction

**DOI:** 10.1101/2021.09.21.21263421

**Authors:** Qijun Gao, Fangfang Bie, Yinfu Hu, Yafeng Chen, Bo Yang

## Abstract

**Background:** At present, the mechanism of reciprocal ST-segment depression(RSTD) is still not clear.

**Methods:** The electrocardiogram and angiography of 85 STEMI patients were retrospectively analyzed to summarize the characteristics of ST segment changes and explore the mechanism of RSTD.

**Results:** A total of 85 patients were included, of which 75 were patients with RSTD (10 patients with anterior myocardial infarction had no RSTD), all 45 patients with inferior myocardial infarction had limb leads RSTD, and 37 of them had anterior lead ST segment depression.Thirty patients with anterior myocardial infarction were accompanied by mild ST segment changes in the limb leads. According to the characteristics of RSTD, it is speculated that the mechanism of RSTD is that the action potential of infarct area decreased, which could not offset the action potential in non-infarct area.

**Conclusion:** the mechanism of RSTD in acute myocardial infarction maybe that the negative electrode action potential of the lead was weakened or disappeared, and the positive electrode action potential could not be completely offset, resulting in ST segment depression.

## Introduction

Acute ST segment elevation myocardial infarction(STEMI) is often accompanied by ST segment depression in the leads opposite to the leads showing ST segment elevation, known as reciprocal ST segment depression (RSTD), which is seen in either ischemia at distance or mirror reflection of ST segment elevation.^[1.2]^ The RSTD mentioned in this paper refers to mirror reflection of ST segment elevation.

At present, RSTD is described as ST segment elevation in any lead that is associated with reciprocal ST segment depression in leads whose positive pole is directed opposite to the leads with ST segment elevation^[3]^. However,the posterior leads do not present RSTD in anterior myocardial infarction, and RSTD in anterior leads also occur in inferior myocardial infarction^[4]^. Therefore, the current view is not accurate, and the mechanisms of RSTD warrant further exploration.

In this study, the electrocardiograms(ECG) of patients with acute STEMI in our hospital were analyzed to summarize the characteristics of RSTD and explore the mechanism of RSTD.

## Patients and Methods

### Study population

The study design is a retrospective analysis of patients admitted to a tertiary hospital (No.1 Hospital of Jingmen City, Hubei Province, China) from January 1, 2018 to March 31, 2021. The study population included patients presenting to the emergency department with symptoms of acute coronary syndrome (ACS) and ST segment elevation in their ECGs on admission.The study was approved by the No.1 Hospital of Jingmen ethics committee and oral consent was obtained from patients.

## Method

All patients subsequently underwent emergency coronary angiographies. The ECG and cardiac catheterization images were retrospectively analyzed by cardiologists.

The diagnosis of STEMI was based on the ECG criteria of ST segment deviations provided by the last consensus document.^[5]^ RSTD was defined as ST segment depression > 0.05 mV in any leads, 40 ms after the J-point.

Exclusion criteria: (1) Patients with non-coronary artery disease or multi-vessel disease (2) Right or left bundle branch block (3)hypertrophic or dilated cardiomyopathy, and (4)significant arrhythmias including atrial fibrillation and ventricular tachycardia.

### Study protocol

Patients were divided into four groups according to their ECG on admission:

1. The anterior STEMI without RSTD; 2. The anterior STEMI with RSTD; 3. Inferior STEMI with RSTD in the anterior lead; 4. Inferior STEMI without RSTD in the anterior lead.

The number of each group and the characteristics of the ST segment changes were summarized.

## Results

A total of 85 patients were included, of which 75 were patients with RSTD (10 patients with anterior myocardial infarction had no RSTD), all 45 patients with inferior myocardial infarction had limb leads RSTD, and 37 of them had anterior lead ST segment depression(Right coronary artery occlusion was found in 28 cases and circumflex artery occlusion in 9 cases).

Thirty patients with anterior myocardial infarction were accompanied by mild ST segment changes in the limb leads. (ST segment depression ≤ 0.1 mV appeared in 25 patients and ST segment depression of 0.1–0.2 mV appeared in 5 patients).

Twenty-five of the patients combined with inferior ST segment depression. The degree of ST segment depression in lead III was approximately equal to the degree of ST segment elevation in lead aVL. The degree of ST segment depression in lead aVF was about half of that in lead III, and there was no significant depression in lead II(Figure 1). There were 5 cases of anterior myocardial infarction with ST segment elevation in inferior leads (less than 0.1 mV) and RSTD in aVR or aVL lead.

**Figure 1.**
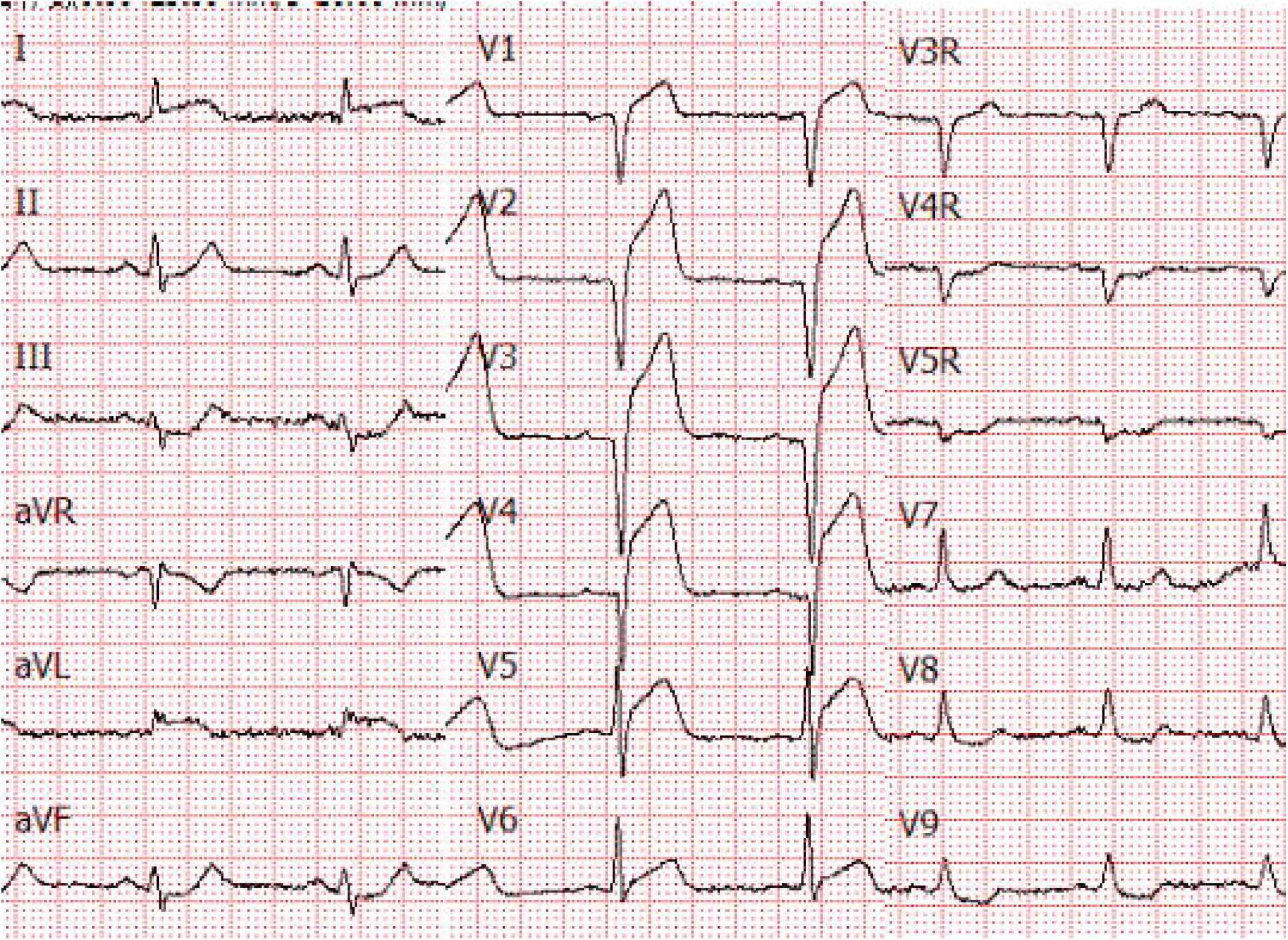
Electrocardiogram of patients with anteriorlateral wall myocardial infarction.

All ECGs of inferior STEMI had aVL or aVR lead ST segment depression. In 34 cases, the degree of ST segment elevation of lead III was greater than that of lead II(Right coronary artery occlusion was found in 26 cases and circumflex artery occlusion in 8 cases). The degree of ST segment depression of lead aVL was significantly greater than that of lead aVR. In two cases, the degree of ST segment elevation of lead II was greater than that of lead III (circumflex artery occlusion), and in 9 cases, the degree of ST segment elevation of lead II was similar to that of lead III(right coronary artery occlusion was found in 1 case and circumflex artery occlusion in 8 cases). The ST segment depression of lead aVL and lead aVR was also similar, which was about half of the elevations of the ST segment in lead III and lead II.

## Discussion

The current view regarding RSTD is not accurate. Specifically, RSTD neither necessarily occurs on the opposite side of the infarct area nor does it occur only on the opposite side of the infarct area. In present study, we noted that RSTD can occur in any lead with the infarct area as the negative pole.

Since the ECG records the potential changes of the positive and negative electrodes^[6]^, the reason for the RSTD should be analyzed from the potential change of the positive and negative electrodes of the leads, respectively.

Transmembrane action potential(AP), formed by transmembrane ions, causes periodic changes in the potential inside myocardial cells, as well as reverse periodic changes in the membrane potential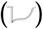. ^[7]^ The membrane potential can be sensed by the electrode. The potential change of positive electrode recorded by the lead is the same as the change in membrane potential 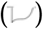, while the potential change of negative electrode is the opposite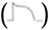,which is similar to the change of intracellular potential.

Under normal conditions, all myocardium changes synchronously,there is no current within the myocardium, and this fluctuation of the extracellular potential is not shown on the ECG. When myocardial injury occurs, the action potential (AP) is weakened, the AP of the normal myocardium cannot be offset by the AP of the injured myocardium, and ST segment changes appear. The ST segment of the lead with the injured area as a positive electrode is elevated, while the ST segment of the lead with the injured area as a negative electrode is depressed.

The relationship between the magnitude of ST-segment elevation and reciprocal ST-segment depression is unclear^[3]^. We found for the first time that the degree of ST-segment depression in limb leads can be calculated from the degree of ST-segment elevation.

The negative electrode of the standard limb leads is a single limb electrode, whereas the negative electrode of the three aV limb leads is the average value of the two limb electrodes, and this must be calculated.^[7]^

When the amplitude of the action potential (AAP) of one limb electrode decreased, the degree of ST segment depressions in the standard limb lead with the infarct area as the negative pole were equal to the degree of ST segment elevation in the lead with the infarct area as the positive pole, while the degree of ST segment depressions in the aV lead were only half of that in the standard limb lead. Figure 1 is an ECG of a patient with acute anterolateral wall STEMI, the ST segment of lead I elevated by 0.1mV, indicated the amplitude of the action potential (AAP) of left arm electrode reduced 0.1mV(assessed from ECG), the left arm electrode is the negative electrode of lead III, half the negative electrode of the lead aVF, so the ST segment depressed 0.1mV in lead III and 0.05mV in the lead aVF.

When two limb electrodes were affected by myocardial infarction, the electrode recording the normal myocardium should be determined first. Figure 3 shows an ECG of a patient with inferior myocardial infarction and right ventricular myocardial infarction, the normal myocardium was recorded by the left arm electrode. The ST segment of lead III elevated by 0.7 mV indicates that the leg electrode decreased by 0.7 mV. The ST segment of lead II elevated by 0.5 mV indicates that the AAP of the leg electrode was 0.5 mV less than that of the right arm electrode,so the AAP of the right arm electrode was 0.2 mV less than that of the left arm electrode. The negative electrode of aVL is the average value of the right arm and leg electrode, AAP decreased 0.45 mV [(0.7 mV + 0.2 mV) / 2], and the ST segment decreased by 0.45 mV. The negative electrode of aVR is the average value of the left arm and leg electrode, AAP decreased 0.35 mV[(0+ 0.7 mV)/2], ST segment decreased by 0.15 mV(0.35 mV -0.2 mV), and lead I decreased by 0.2 mV.

**Figure 2.**
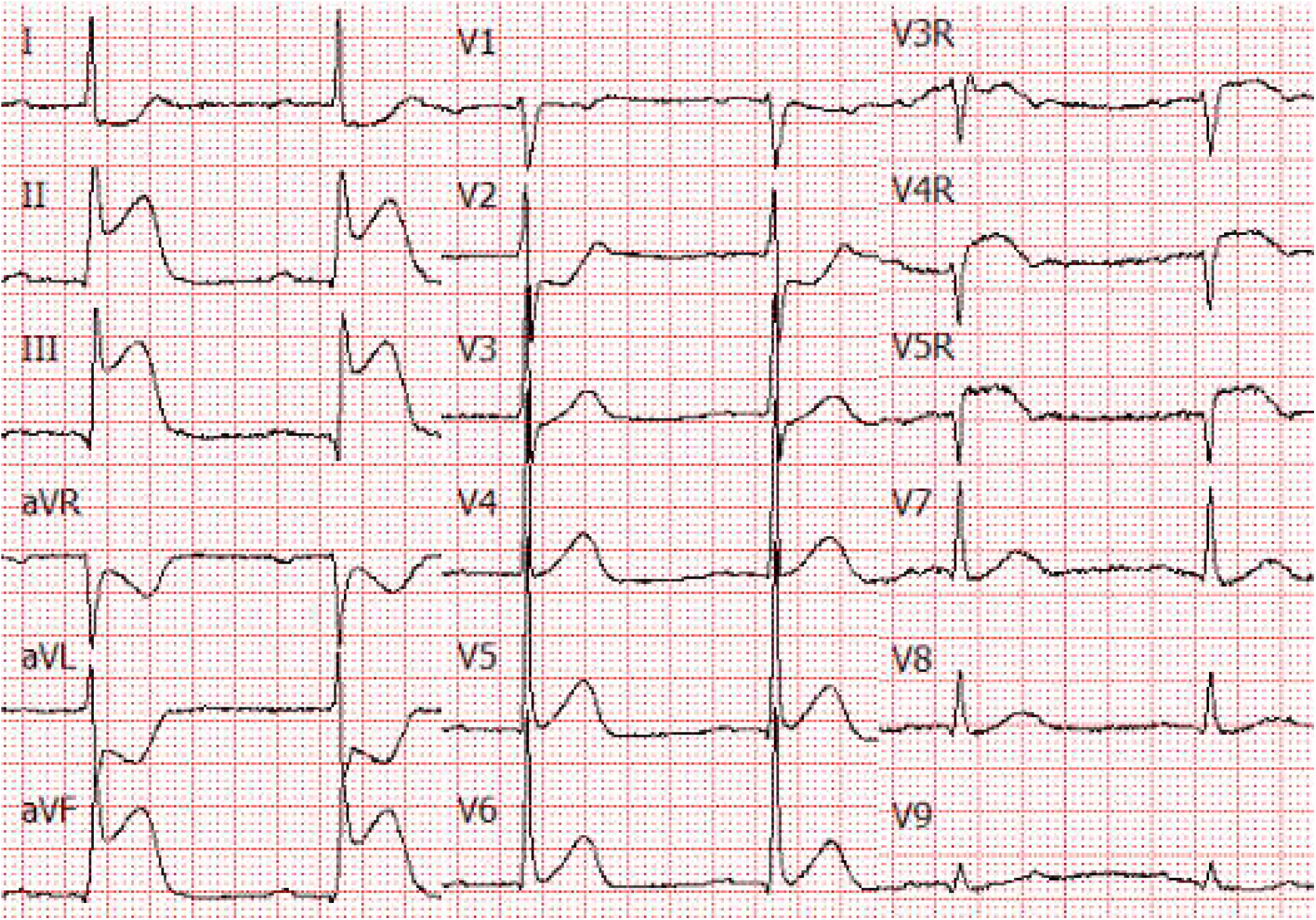
Electrocardiogram of a patient with acute inferior wall ST segment elevation myocardial infarction and right ventricular myocardial infarction.

**Figure 3.**
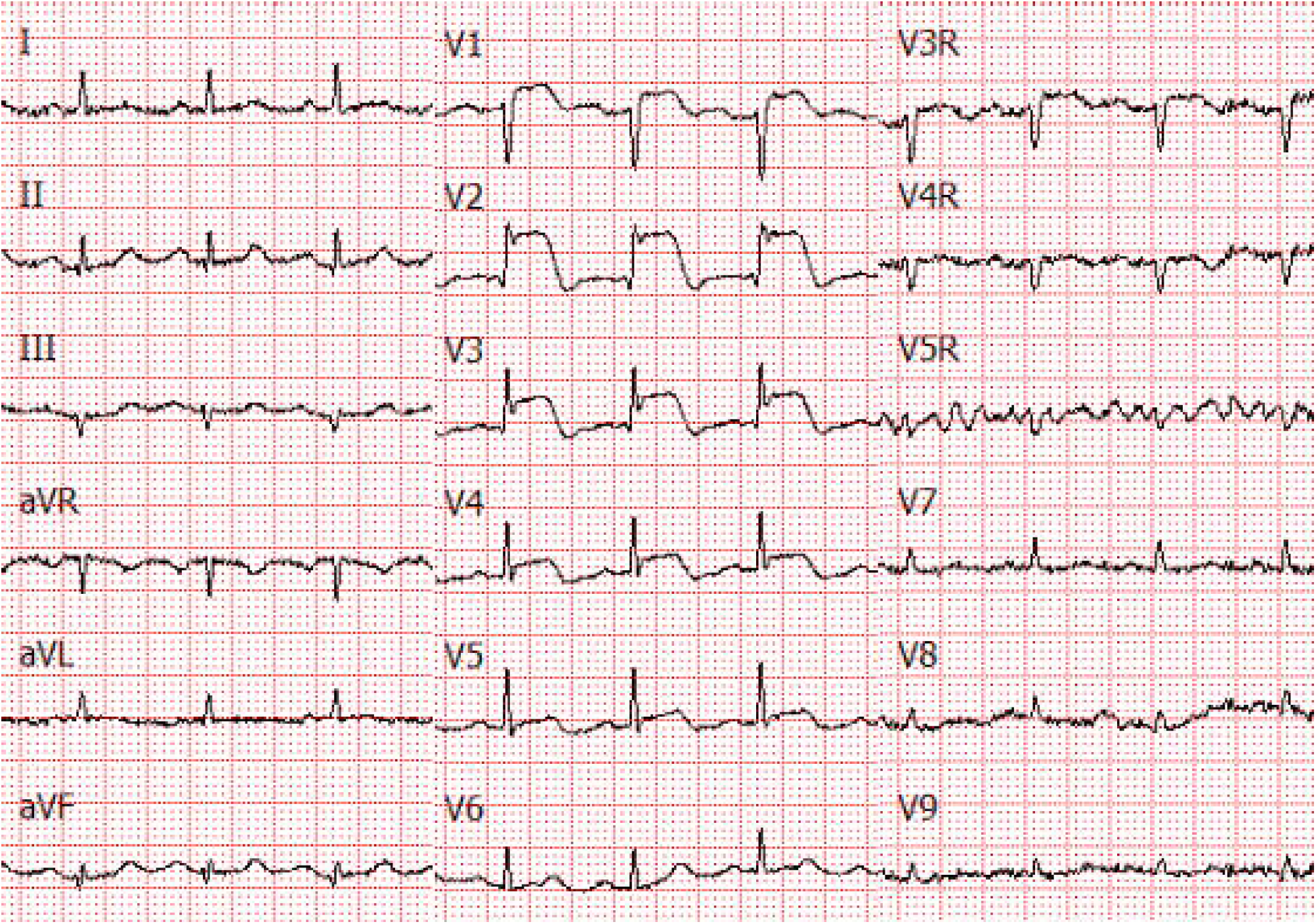
Electrocardiogram of a patient with acute anterior ST segment elevation myocardial infarction.

According to the current view, anterior wall myocardial infarction should have RSTD in the posterior wall^[3]^, but this is not the case (Figure 3). This is because the negative pole of all leads is in the limb leads, and part of the anterior wall myocardial infarction may not involve the limb leads. In this study, there were 10 patients with anterior myocardial infarction who showed no RSTD in any leads. There were 25 patients with anterior wall myocardial infarction combined with left lateral wall myocardial infarction and 5 patients with inferior wall myocardial infarction. The range of the inferior wall or left lateral wall myocardial infarction in patients with anterior wall myocardial infarction is generally small, with slight ST segment elevation in limb leads and most patients do not have RSTD in the posterior leads.

Currently the depression in leads V1and V2 is considered the reciprocal of and similar in meaning to the ST-segment elevation in the V8 and V9.^[3]^ However, depression in leads V1and V2 is also seen in right ventricular myocardial infarction. ^[8]^ The negative electrode of precordial leads is the Wilson’s Central Terminal, which is the average value of three limb electrodes. Myocardial infarcts with ST segment elevation in limb leads may cause RSTD in precordial leads. The depression in leads V1 and V2 is the RSTD of ST elevation in the inferior leads, not the posterior or right ventricular leads. The ST segment depression of V1 was rare in right ventricular myocardial infarction, because right ventricular action potential recorded by V1 was decreased too. ST segment was mostly normal or elevated and only a few showed depression.

The reason for the absence of RSTD in the precordial leads in inferior infarction was that the precordial electrode was close to the infarct area and the AAP was also reduced. Another reason was that the AAP of the inferior myocardium was slightly weakened, which had no obvious effect on the potential of the negative electrode of the precordial lead. Among the 13 patients with inferior myocardial infarction without ST segment depression in the anterior leads, ST segment elevation in the inferior leads was 0.1 mV in five patients, 0.1–0.2 mV in seven patients, and 0.4 mV in one patient(combined with extensive anterior wall ST segment elevation).

The link between RSTD and clinical outcomes has been extensively studied since long, and majority of the previous research suggests that RSTD is associated with clinical prognosis.^[9-14]^ ST segment depression occurs in some patients due to multivessel lesions, and the prognosis of these patients is poor. However, even in patients without multivessel disease, RSTD may be associated with poor prognosis. RSTD is related to the degree of ST segment elevation—the more the ST segment elevation in limb leads, the greater the risk of ST segment depression in chest leads. Furthermore, the degree of RSTD between limb leads is directly related to the degree of ST segment elevation in the infarct area.

## Limitations

It is difficult to obtain patients’ local myocardial action potential clinically. Thus, we can only speculate about the mechanism in terms of the characteristics of ST segment change, but the mechanism we have proposed completely conforms to the characteristics of ST segment change, making it the most reasonable and scientific mechanism so far. It is expected that experiments on animals could obtain the local action potential of the myocardium and directly verify the proposed mechanism.

In summary, the mechanism of RSTD in acute myocardial infarction maybe that the AP of negative electrode of the lead was weakened or disappeared, and the AP of positive electrode could not be completely offset, resulting in ST segment depression.

## Data Availability

All information may be obtained by contacting the author of the newsletter

https://m0.mail.sina.com.cn/classic/index.php#action=maillist&fid=unk&title&pagecount=0&pageno=1&order=htime&sorttype=desc&type=0&tag=-1

## Contributorship Statement

The study was designed by QG and BY, FB collected the ECG,QG drafted the manuscript, YH contributed to critical revision.

